# APOE ε4 Carrier Status and Cognitive Decline in Mild-to-Moderate Alzheimer’s Disease: A Linear Mixed-Effects Analysis of the Placebo Arm of EXPEDITION 1

**DOI:** 10.64898/2026.09.27.26364131

**Authors:** Amirhomayoun Heidari, Mitra Aghajani

## Abstract

**Background:** Studies indicate that the apolipoprotein E (APOE) ε4 allele is the strongest genetic risk factor for Alzheimer’s disease (AD), and individuals with this allele are at increased risk of developing AD, there remains debate regarding whether APOE ε4 carriers experience more rapid cognitive decline over time compared to non-carriers. This study assessed this question using linear mixed-effects models applied to the placebo arm of EXPEDITION 1, a phase 3 clinical trial evaluating solanezumab in patients with mild-to-moderate Alzheimer’s disease.

**Methods:** In this study we utilized data from 370 participants in the placebo group of the EXPEDITION 1 trial. After excluding 27 participants with undefined APOE genotype, all models and analyses were applied to the remaining 343 participants (686 ADAS-Cog14 observations) to determine whether APOE ε4 carriers exhibit a faster rate of decline over the 80-week follow-up period. Five nested linear mixed-effects models with a participant-specific random intercept were fitted, and also a complementary change-score analysis was done.

**Results:** Over 80 weeks, ADAS-Cog14 worsened by an average of 7.01 points. Carriers did not decline faster than non-carriers (additional decline 1.31 points; 95% CI −1.22 to 3.83; p = 0.310), and the change-score analysis showed the same pattern. Carriers had worse ADAS-Cog14 scores overall, but this difference was no longer significant after adjustment for baseline CDR-SB.

**Conclusion:** In this cohort, APOE ε4 carrier status was related to disease severity at baseline rather than to the rate of cognitive decline over 80 weeks.

## Introduction

Alzheimer’s disease (AD) is the most common cause of dementia, responsible for 60-70 percent of cases, and in wealthy nations, it ranks among the leading causes of death. Pathologically, it is defined by the buildup of amyloid beta (Aβ) protein fragments and clustering of tau protein that forms tangles inside neurons.^1, 2^

The apolipoprotein E (APOE) ε4 allele is recognized as the strongest genetic risk factor for AD. Studies showed that carrying the ε4 allele increases the risk of developing AD.^3^

Individuals who carry one ε4 allele have a significantly higher risk of developing AD in comparison to the non-carriers, while those with two ε4 alleles show an even greater increase in risk. In contrast, the ε2 allele appears to have a protective effect. Although the strength of the association varies across ethnic groups, APOE ε4 has consistently been associated with increased AD susceptibility across different populations, ages, and both sexes.^4^

Studies have also shown an association between APOE ε4 and cognitive decline.^5, 6^ In a large autopsy-confirmed cohort, results showed that the rate of Clinical Dementia Rating Sum of Boxes (CDR-SB) increase in the APOE ε4 carriers was approximately 1.5 times faster than in APOE ε3/ε3 carriers and about 1.3 times faster than in APOE ε2 carriers. APOE ε4 carriers also showed about 1.1 times faster Mini-Mental State Examination (MMSE) decline compared with APOE ε3/ε3 carriers. In contrast, APOE ε2 carriers showed a slower MMSE decline than APOE ε3/ε3 carriers, but their difference in CDR-SB progression was not statistically significant.^7^

Previous studies examining the association between APOE ε4 and the rate of cognitive decline have reported inconsistent findings, which may partly reflect differences in disease stage and statistical modeling approaches.^2, 8^ For example, one study of 649 participants with biomarker-confirmed early-stage AD assessed cognitive decline using MMSE, CDR-SB, and the 13-item Alzheimer’s Disease Assessment Scale–Cognitive Subscale (ADAS-Cog13) and found no significant difference in the overall rate of cognitive decline between APOE ε4 carriers and non-carriers. However, the effect varied by disease stage: ε4 carriers with late mild cognitive impairment showed a faster decline in MMSE, whereas ε4 carriers with mild AD showed a slower decline in MMSE and CDR-SB compared with non-carriers.^2^ The relationship between APOE ε4 and the rate of cognitive decline remains unclear and controversial, particularly in well-characterized populations with standardized longitudinal follow-up. Phase 3 trials of solanezumab for mild-to-moderate Alzheimer’s disease, including EXPEDITION 1 and 2, did not demonstrate significant improvement in primary outcomes. Cognitive and functional abilities did not differ significantly between the solanezumab and placebo groups.^9^

A subsequent trial (EXPEDITION 3) involving 2129 participants with mild dementia due to Alzheimer’s disease divided participants into solanezumab (n=1057) and placebo (n=1072) groups. The difference in cognitive decline between the solanezumab and the placebo groups was not statistically significant.^10^ Although these studies did not meet the initial endpoints, they have provided well-characterized longitudinal datasets.

Cognitive decline can vary considerably among patients with Alzheimer’s disease, even when they do not receive treatment. In the EXPEDITION trial, 46.3% of patients in the placebo group showed no cognitive decline over 18 months, and a similar proportion was observed in EXPEDITION 2 (45.6%).^11^ This suggests that patients with Alzheimer’s disease do not all decline at the same rate. However, it is still unclear whether factors such as APOE ε4 status help explain these differences in cognitive decline.

The objective of this study was to examine whether being APOE ε4 carrier is associated with faster cognitive decline over a period of 80 weeks in mild-to-moderate AD patients. Mixed-effects models were applied to the placebo arm of the EXPEDITION 1 trial to evaluate the hypothesis of this study. Our hypothesis was that ε4 carriers would exhibit faster cognitive decline (ADAS-Cog14) in comparison to the non-carrier group.

## Methods

### Study design and data source

This study is an observational secondary analysis using patient-level data from EXPEDITION 1, a Phase 3, randomized, double-blind, placebo-controlled trial of intravenous solanezumab in people with mild-to-moderate Alzheimer’s disease (AD). EXPEDITION 1 was registered at Clinical Trials.gov number (NCT00905372). The original trial protocol was approved by the institutional review board at each participating site, and written informed consent was obtained from all participants. The trial included 1,012 patients, aged 55 years or older without clinically significant depression, and they were followed for 80 weeks; however, neither of the co-primary endpoints was met.^9^ While treatment was randomized in the original trial, the exposure in this analysis, apolipoprotein E (APOE) ε4 carrier status, was not.

The dataset used for this analysis included 2,223 observations of 18 different variables from 741 patients, each with records from three visits at different time points: screening (VISID 1), baseline (VISID 2), and the final follow-up at week 80 (VISID 23).^9^ The Alzheimer’s Disease Assessment Scale–Cognitive subscale, 14-item version (ADAS-Cog14), with a range of 0 to 90, where higher scores indicate worse cognition, was measured at baseline and week 80.^12, 13^ The Mini-Mental State Examination (MMSE) was administered at each visit.^14^ The Geriatric Depression Scale (GDS) was assessed only at screening.^15^ Depression was considered absent with a GDS score of 6 or lower (range 0 to 15, with higher scores indicating greater severity).^9^

The Clinical Dementia Rating–Sum of Boxes (CDR-SB)^16^, the Alzheimer’s Disease Cooperative Study– Activities of Daily Living scale (ADCS-ADL)^17^, and hippocampal volumes were also measured at baseline and week 80.^9^ Ventricular and whole-brain volumes were recorded only at baseline. Higher ADAS-Cog14 and CDR-SB scores indicate worsening condition in the patient, while lower MMSE and ADCS-ADL scores also reflect decline. Therefore, cognitive decline is shown by an increase in ADAS-Cog14 score.

### Derivation of the exposure

APOE ε4 carrier status was derived from the recorded APOE genotype by pattern matching on the genotype string: any participant in this dataset whose genotype contained at least one ε4 allele (E2/E4, E3/E4, or E4/E4) was classified as a carrier, and participants whose genotype contained no ε4 allele (E2/E2, E2/E3, or E3/E3) were considered as non-carriers, and participants with a blank genotype field as missing. Out of 741 participants, the APOE genotypes were distributed as follows: E2/E2 (1 person), E2/E3 (n=32), E2/E4 (n=17), E3/E3 (n=227), E3/E4 (n=323), and E4/E4 (n=91). The genotype information was missing for the other 50 participants. This classification yielded 431 carriers, 260 non-carriers, and 50 participants with unknown status in the full dataset.

### Construction of the baseline record and the longitudinal dataset

For ease of further analysis, we generated a baseline record for each participant. Baseline MMSE was calculated as the average of screening and baseline measurements, or as the available value when only one was present. GDS, recorded only at screening, served as the baseline GDS. All other baseline variables, including ADAS-Cog14, ADCS-ADL, CDR-SB, age, sex, education, treatment, race, ethnicity, and MRI volumes, were obtained from the baseline visit (VISID 2).

The final longitudinal dataset used for the mixed-effects analyses contained two records per participant: one for VISID 2 and one for VISID 23 (week 80), with ADAS-Cog14 as the outcome. Baseline GDS and baseline CDR-SB were included as time-fixed covariates, so CDR-SB reflected baseline severity rather than changes over time.

### Analysis population

We limited our analyses to the placebo group to ensure that any observed cognitive decline was not attributable to the study drug. Because of this, treatment assignment is not included in any model. Out of 370 participants in the placebo group, we excluded 27 who did not have known APOE ε4 carrier status.

This left us with 343 participants at baseline (119 non-carriers and 224 carriers) and a total of 686 observations across two visits. All 343 participants of the study had complete ADAS-Cog14 data at both visits (Figure 1).

**Figure 1.**
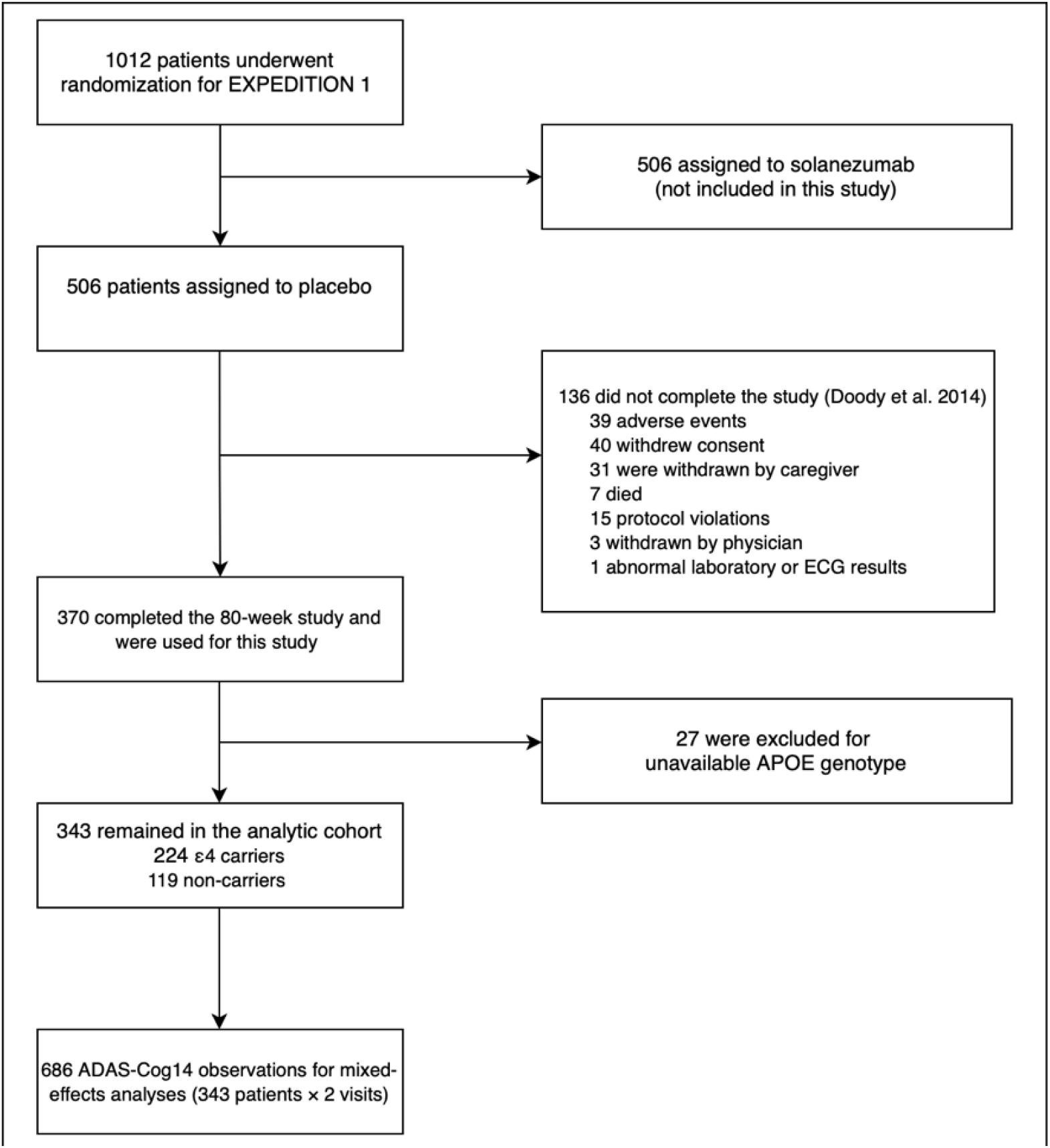
Flow of participants. Randomization, allocation, and discontinuation counts (n = 1,012, 506, and 136) are from the published trial report.^9^ All subsequent counts were produced from the shared patient-level dataset.

Prior to the modeling, the longitudinal dataset was limited to observations with complete data for the outcome, time, and all model covariates (ADAS-Cog14, VISID, carrier status, age, sex, education, GDS, and CDR-SB). This approach ensured that a single complete-case dataset was used for all models, allowing for direct comparison of information criteria. As baseline GDS and baseline CDR-SB were complete for all 686 longitudinal observations, no data were excluded, resulting in a complete-case dataset identical to the full longitudinal analytical population (686 observations from 343 participants). Some baseline imaging variables were incomplete: left and right hippocampal volumes were available for 326 of 343 participants, and ventricular and whole-brain volumes for 332 of 343 participants. These variables are reported with their respective denominators.

## Statistical analysis

The distributions of eleven continuous baseline variables were examined with histograms, boxplots, and Shapiro–Wilk tests.^18^ After examination, all Shapiro–Wilk tests showed non-normality at the 0.05 level (W ranged from 0.8800 to 0.9908). Since normality tests can be too sensitive with larger samples, we mainly relied on the graphical distributions to choose summary statistics and hypothesis tests for the data. For those variables which looked roughly symmetric (age, education, MMSE, ADAS-Cog14, left and right hippocampal volume, whole-brain volume), we reported the mean and standard deviation (SD) and used Welch two-sample t-tests for comparisons, and for skewed variables (CDR-SB, ADCS-ADL, GDS, ventricular volume), we reported the median and interquartile range and used Mann–Whitney U tests.^19-23^ In the placebo arm, baseline characteristics were compared between ε4 carriers and non-carriers to generate Table 1. Symmetric continuous variables were analyzed using the Welch two-sample t-test, while skewed variables were evaluated with the Wilcoxon rank-sum test (Mann–Whitney U) with continuity correction.^24-28^ For categorical variables, Pearson’s chi-squared test was applied when all expected cell counts were at least 5; otherwise, Fisher’s exact test was used. Sex was compared using the chi-squared test with Yates’ continuity correction for 2 × 2 tables, and race, which included several categories with small, expected counts, was analyzed using Fisher’s exact test. Percentages for categorical variables were calculated within each carrier group among participants without missing values.^29-33^

**Table 1.** Baseline characteristics of the placebo-arm analytical population by APOE ε4 carrier status.

| Characteristic | Non-carrier (n=119) | Carrier (n=224) | p-value |
| --- | --- | --- | --- |
| Age, years | 74.7 ± 9.0 | 74.5 ± 7.2 | 0.790 |
| Education, years | 12.5 ± 4.5 | 12.8 ± 3.8 | 0.521 |
| MMSE | 21.5 ± 2.9 | 21.1 ± 2.9 | 0.197 |
| ADAS-Cog14 | 30.7 ± 10.4 | 33.2 ± 9.7 | 0.033 |
| CDR-SB | 4.0 [3.0–6.0] | 4.8 [3.5–6.0] | 0.098 |
| ADCS-ADL | 65.0 [54.0–71.0] | 65.0 [56.8–71.0] | 0.558 |
| GDS | 2.0 [1.0–3.0] | 1.0 [1.0–2.0] | 0.287 |
| Left hippocampus | 1856.7 ± 405.1 | 1637.6 ± 358.7 | <0.001 |
| Right hippocampus | 1892.0 ± 421.4 | 1694.9 ± 360.5 | <0.001 |
| Ventricular volume | 43.3 [30.7–66.1] | 45.5 [31.8–58.8] | 0.815 |
| Whole-brain volume | 1004.5 ± 110.0 | 1006.6 ± 106.9 | 0.866 |
| Sex, n(%) |  |  | 0.176 |
| F | 63 (52.9) | 137 (61.2) |  |
| M | 56 (47.1) | 87 (38.8) |  |
| Race, n(%) |  |  | 0.980 |
| American Indian or Alaska Native | 0 (0.0) | 1 (0.4) |  |
| Asian | 14 (11.8) | 27 (12.1) |  |
| Black or African American | 4 (3.4) | 7 (3.1) |  |
| Multiple | 0 (0.0) | 2 (0.9) |  |
| White | 101 (84.9) | 187 (83.5) |  |

Symmetric variables are mean ± SD (Welch two-sample t-test); skewed variables are median [Q1–Q3] (Wilcoxon rank-sum test with continuity correction). Hippocampal volumes were available only for 326 of 343 participants (110 non-carriers, 216 carriers). Ventricular and whole-brain volumes were also available for 332 participants (113 non-carriers, 219 carriers). Sex was compared using Pearson's chi-squared test with Yates' continuity correction, and race using Fisher's exact test.

The longitudinal changes in ADAS-Cog14 were analyzed using linear mixed-effects models^34^ constructed from the complete-case longitudinal dataset mentioned above (which consisted of 686 observations from 343 participants), the parameters being estimated by maximum likelihood (as opposed to restricted maximum likelihood, in order to enable likelihood-ratio tests between models that differed in their fixed effects). In these models a participant-specific random intercept was included to consider the correlation between the two repeated measurements made on each participant. Time was expressed in weeks of follow-up, taking the value 0 at baseline and 80 at the final visit. All coefficients relating to time are given per week and, where indicated, rescaled to the 80-week period by multiplying by 80. Tests of the fixed effects were carried out using Satterthwaite’s approach for the degrees of freedom, and 95% confidence intervals (CIs) for all the fixed effects were computed using the Wald method.

Five models were fitted in turn, each one aiming to answer a particular research question. The first model contained time, age, sex and education to establish the average trajectory of ADAS-Cog14 after allowing for the effects of the basic demographic variables. The second model added APOE ε4 carrier status to find out if carriers differed from non-carriers in their overall ADAS-Cog14 scores after taking the demographic factors into account. Model 3 included baseline GDS so that it could be examined whether depressive symptoms accounted for any of the differences observed between carriers and non-carriers. Model 4 incorporated baseline CDR-SB to assess whether the difference in ADAS-Cog14 levels between carriers and non-carriers remained after adjustment for the general severity of dementia. Model 5 introduced the interaction between time and APOE ε4 carrier status, and this was the main hypothesis test to see whether the rate of change in ADAS-Cog14 over the 80-week period was different for carriers and non-carriers. The main effect of APOE in Models 2 to 5 refers to a difference in ADAS-Cog14 levels between carriers and non-carriers, not a difference in the rate of decline, and is given as a secondary finding. Successive models were compared using likelihood-ratio tests and the Akaike information criterion (AIC).^5, 6^

As a complementary analysis, the ADAS-Cog14 change score for each participant was computed as the week-80 score minus the baseline score, with positive values indicating greater cognitive decline. Change scores were analyzed using linear regression within the same complete-case cohort of 343 participants utilized in the mixed-effects models. Models were constructed incrementally: the first adjusted for age, sex, and education; the second included APOE ε4 carrier status; and the third incorporated baseline GDS. Since ADAS-Cog14 was measured at two time points, both the change-score analysis and the time **×** carrier interaction in the mixed-effects models address the same research question.

All analyses were performed in R version 4.3.3^35^. Mixed-effects models were fitted with lme4 (version 1.1-37)^36^ with Satterthwaite inference from lmerTest (version 3.1-3)^37^.

## Results

### Analysis population and baseline characteristics

The shared dataset included 741 participants, who all completed the 80-week follow-up period. A total of 370 participants received a placebo, but 27 were removed later because we did not have their APOE genotype data. This resulted in a final analysis group of 343 patients, providing 686 total ADAS-Cog14 scores over time. In this group of 343 participants, 224 (65.3%) had at least one ε4 allele, and 119 (34.7%) did not carry any ε4 allele.

Baseline characteristics by carrier status are presented in Table 1. Carriers and non-carriers were comparable in age (74.5 ± 7.2 vs 74.7 ± 9.0 years; p = 0.790), sex distribution (61.2% vs 52.9% female; p = 0.176), education (12.8 ± 3.8 vs 12.5 ± 4.5 years; p = 0.521), race (p = 0.980), and baseline MMSE scores (21.1 ± 2.9 vs 21.5 ± 2.9; p = 0.197). Carriers showed higher baseline ADAS-Cog14 scores than non-carriers (33.2 ± 9.7 vs 30.7 ± 10.4; p = 0.033), reflecting poorer baseline cognition, as well as smaller left (1637.6 ± 358.7 vs 1856.7 ± 405.1 mm^3^ ; p < 0.001) and right (1694.9 ± 360.5 vs 1892.0 ± 421.4 mm^3^ ; p < 0.001) hippocampal volumes (data available for 326 of 343 participants). Median [interquartile range] CDR-SB was 4.8 [3.5–6.0] in carriers and 4.0 [3.0–6.0] in non-carriers (p = 0.098), while median GDS was 1.0 [1.0–2.0] and 2.0 [1.0–3.0], respectively (p = 0.287). No significant differences were observed between groups for ADCS-ADL (p = 0.558), ventricular volume (p = 0.815; n = 332), or whole-brain volume (p = 0.866; n = 332).

### Longitudinal models

Table 2 illustrates the results of the five mixed-effects models made in this study. All models used the same complete-case dataset with 686 observations of 343 participants.

**Table 2.**
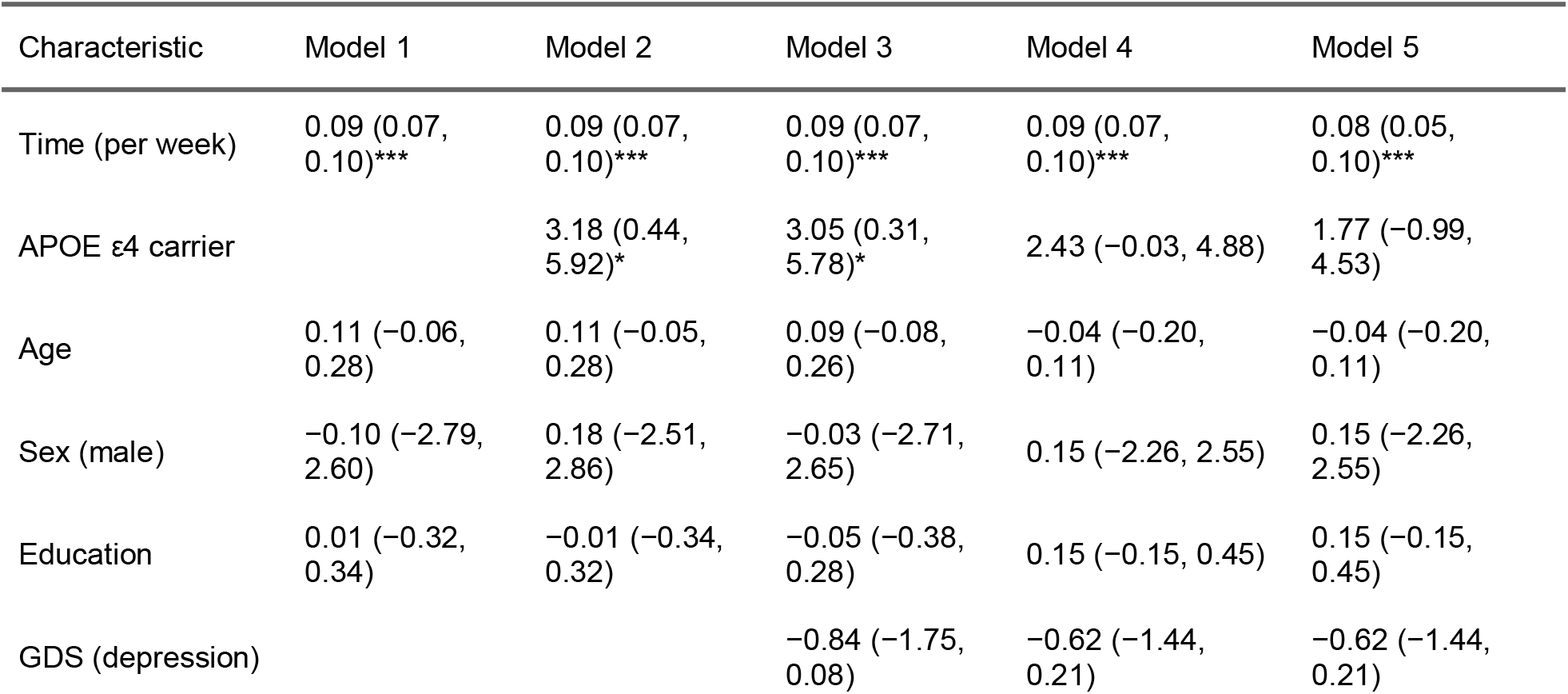
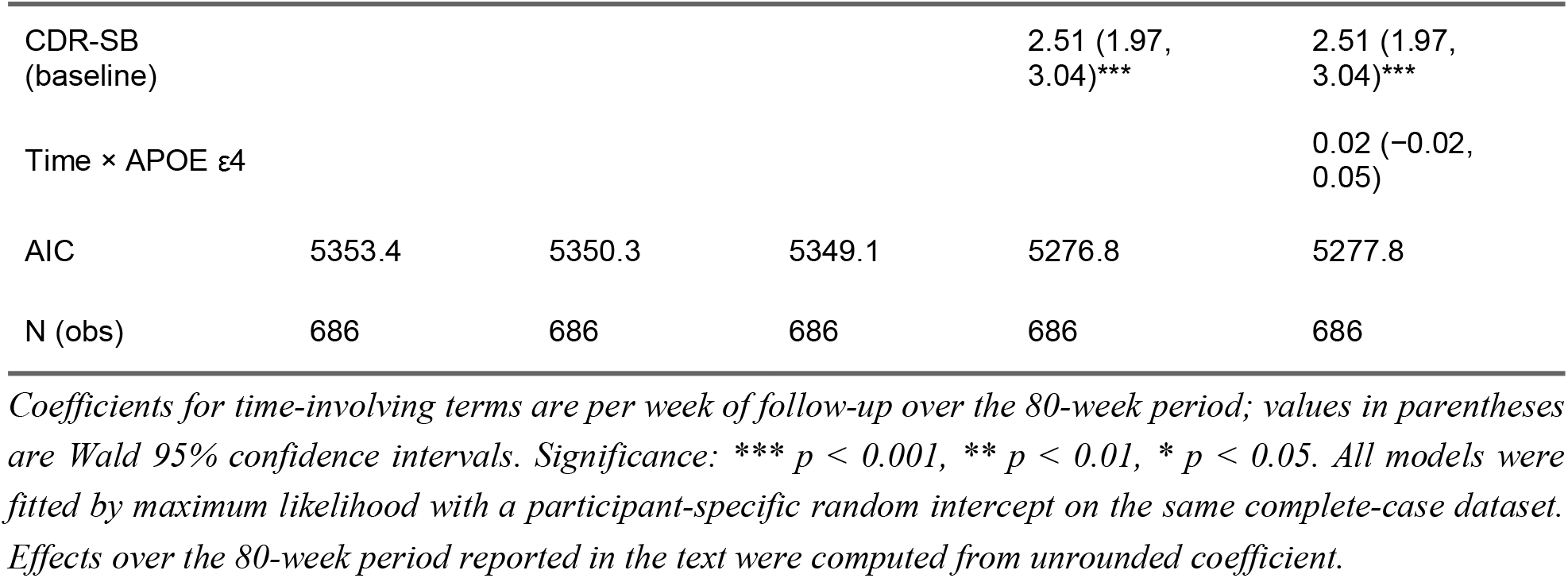
Linear mixed-effects models for ADAS-Cog14 (686 observations, 343 participants in placebo arm).

| Characteristic | Model 1 | Model 2 | Model 3 | Model 4 | Model 5 |
| --- | --- | --- | --- | --- | --- |
| Time (per week) | 0.09 (0.07, 0.10)*** | 0.09 (0.07, 0.10)*** | 0.09 (0.07, 0.10)*** | 0.09 (0.07, 0.10)*** | 0.08 (0.05, 0.10)*** |
| APOE $\epsilon 4$ carrier | | 3.18 (0.44, 5.92)* | 3.05 (0.31, 5.78)* | 2.43 ($-0.03$, 4.88) | 1.77 ($-0.99$, 4.53) |
| Age | 0.11 ($-0.06$, 0.28) | 0.11 ($-0.05$, 0.28) | 0.09 ($-0.08$, 0.26) | $-0.04$ ($-0.20$, 0.11) | $-0.04$ ($-0.20$, 0.11) |
| Sex (male) | $-0.10$ ($-2.79$, 2.60) | 0.18 ($-2.51$, 2.86) | $-0.03$ ($-2.71$, 2.65) | 0.15 ($-2.26$, 2.55) | 0.15 ($-2.26$, 2.55) |
| Education | 0.01 ($-0.32$, 0.34) | $-0.01$ ($-0.34$, 0.32) | $-0.05$ ($-0.38$, 0.28) | 0.15 ($-0.15$, 0.45) | 0.15 ($-0.15$, 0.45) |
| GDS (depression) | | | $-0.84$ ($-1.75$, 0.08) | $-0.62$ ($-1.44$, 0.21) | $-0.62$ ($-1.44$, 0.21) |
| CDR-SB (baseline) |  |  |  | 2.51 (1.97, 3.04)*** | 2.51 (1.97, 3.04)*** |
| Time × APOE ε4 |  |  |  |  | 0.02 (−0.02, 0.05) |
| AIC | 5353.4 | 5350.3 | 5349.1 | 5276.8 | 5277.8 |
| N (obs) | 686 | 686 | 686 | 686 | 686 |

*Coefficients for time-involving terms are per week of follow-up over the 80-week period; values in parentheses are Wald 95% confidence intervals. Significance: \*\*\* $p < 0.001$ , \*\* $p < 0.01$ , \* $p < 0.05$ . All models were fitted by maximum likelihood with a participant-specific random intercept on the same complete-case dataset. Effects over the 80-week period reported in the text were computed from unrounded coefficient.*

In Model 1, ADAS-Cog14 increased by 0.09 points per week (95% CI 0.07 to 0.10 and p < 0.001), corresponding to an average worsening of 7.01 points (95% CI 5.80 to 8.21) over the 80-week period and the results showed that age, sex and education were not associated with ADAS-Cog14.

In Model 2, ε4 carriers scored on average 3.18 points higher (worse) than non-carriers across the study period (95% CI 0.44 to 5.92; p = 0.024). The likelihood-ratio test for adding carrier status was χ^2^(1) = 5.14, p = 0.023.

In Model 3, additional adjustment for baseline GDS did not alter the carrier difference (3.05 points; 95% CI 0.31 to 5.78; p = 0.030), and GDS itself was not associated with ADAS-Cog14 at the 0.05 level (−0.84 per point; 95% CI −1.75 to 0.08; p = 0.074).

When baseline CDR-SB was considered in Model 4, the results changed substantially. CDR-SB was strongly associated with ADAS-Cog14 (2.51 points per CDR-SB point; 95% CI 1.97 to 3.04; p < 0.001; likelihood-ratio χ^2^(1) = 74.33), and the AIC decreased from 5349.1 to 5276.8. With CDR-SB in the model, the APOE ε4 carrier difference decreased from 3.05 to 2.43 points and was no longer statistically significant (95% CI −0.03 to 4.88; p = 0.053).

The main question of this study was whether APOE ε4 carriers declined faster than non-carriers over 80 weeks or not, and Model 5 tested this question. The time × APOE ε4 interaction was 0.02 points per week (95% CI −0.02 to 0.05). Because the interval includes zero, no significant difference in the rate of decline was found. Over the full 80-week period, this corresponded to an additional 1.31 points in carriers (95% CI −1.22 to 3.83; p = 0.310; likelihood-ratio χ^2^(1) = 1.03). Non-carriers worsened by 0.08 points per week (95% CI 0.05 to 0.10; p < 0.001), and carriers scored 1.77 points higher at baseline (95% CI −0.99 to 4.53), a difference that was also not significant. Adding the interaction did not improve model fit (AIC 5277.8 vs 5276.8 in Model.

As a check, we repeated the analysis using change scores and obtained the same result. In the change-score analysis, the mean ADAS-Cog14 change over the 80-week period was 7.01 points. After adjustment for age, sex, and education, the difference in change between ε4 carriers and non-carriers was 1.23 points (95% CI −1.30 to 3.76; p = 0.339), which was nearly identical to the estimate from the mixed-effects interaction model (1.31 points; 95% CI −1.22 to 3.83). Further adjustment for baseline GDS did not materially alter the estimate (1.18 points; 95% CI −1.36 to 3.72).

## Discussion

In this study on the placebo arm of EXPEDITION 1, ε4 carriers did not exhibit faster cognitive decline over 80 weeks (1.31 additional points; 95% CI −1.22 to 3.83), and the change-score analysis gave a similar estimate (1.23 points). ε4 carriers on average showed worse ADAS-Cog14 scores (3.18 points higher), but after adjustment for baseline CDR-SB the difference became 2.43 points and was not significant anymore.

Our null result is in contrast with a study of 1,102 autopsy-confirmed AD cases, which tested whether APOE genotype is associated with differences in decline rate among patients. That study reports that CDR-SB increases at a rate 1.5 times faster in APOE ε4 carriers than in APOE ε3/ε3 carriers, and approximately 1.3 times faster than in APOE ε2 carriers,^7^ and another study that reported faster cognitive decline in ε4 carriers over approximately four years of follow-up with five visits.^3^

But our findings are consistent with a study reporting no significant difference in the overall rate of cognitive decline among 649 biomarker-confirmed early Alzheimer’s disease participants. The study reported that the APOE ε4 allele appears to have minimal impact on cognitive decline rates in the study cohort.^2^ It is important to consider that in our study we had a shorter follow-up of 80 weeks with only two assessments, which may not have been enough to detect small differences in decline rate.

Our results showed that CDR-SB was strongly associated with ADAS-Cog14 (2.51 points per point) and its inclusion reduced the AIC by more than 70 points and made the carrier difference non-significant. This may suggest that carriers’ apparent disadvantage largely reflected entering the study at a more advanced stage which is consistent with their worse baseline ADAS-Cog14 and smaller hippocampi, as shown in Table 1. In line with this, a large study of APOE ε4 homozygotes reported that almost all of them developed AD pathology and showed symptom onset at an earlier and highly predictable age, supporting the idea that ε4 mainly influences when the disease begins.^38^ One caution is that CDR-SB itself partly measures cognition, so adjusting for it may remove some of the true carrier effect as well.

## Limitations

This study has limitations. The dataset contained only participants who completed the study (136 of 506 placebo participants discontinued), and preferential loss of faster-declining carriers would bias results toward the null. With two assessments we could estimate net change but not trajectory shape, and 80 weeks may be too short for slope differences to emerge. Because the exposure was observational, we cannot draw any conclusions about causality. Carrier status was treated as a binary variable, and we did not look at allele-dose effects.

In conclusion, findings from this study and 80 weeks of follow-up on the placebo arm of EXPEDITION 1 suggest that ε4 status may indicate patient’s position within the disease trajectory rather than the rate of cognitive decline. With extended follow-up, more frequent assessments, and consideration of allele dosage, it may be possible to clarify whether the presence of the ε4 allele influences the speed of cognitive decline over time.

## Data Availability

All data produced in the present study are available upon reasonable request to the authors

## Acknowledgement

This secondary analysis received no specific funding. The authors declare no conflicts of interest. The author reviewed and approved all content and takes full responsibility for it.

